# Exploring the genetic determinants of ALS in a North Indian cohort using a genomic screening approach

**DOI:** 10.64898/2026.11.20.26368383

**Authors:** Saileyee Roychowdhury, Deepika Joshi, Anand Kumar, Parimal Das

**Affiliations:** Centre for Genetic Disorders, Institute of Science, Banaras Hindu University, Varanasi, India; Department of Neurology, Institute of Medical Sciences, Banaras Hindu University, Varanasi, India

**Keywords:** ALS genetics, Indian population, Sleep disturbances, *NEK1*, *TARDBP*, *SETX*, *ERBB4*, *ANXA11*, *SIGMAR1*

## Abstract

**Background:** Amyotrophic lateral sclerosis is a clinically and genetically heterogenous neurodegenerative disorder in which both monogenic and complex genetic mechanism contribute to disease susceptibility. Increasing evidence also indicates that non motor manifestations, including sleep disturbances, significantly influence disease burden. This study aimed to characterize the genetic spectrum of ALS in a predominantly North Indian cohort while evaluating the prevalence of sleep related abnormalities.

**Methods:** Seventy-three clinically diagnosed ALS patients were recruited from a tertiary care centre in North India. Disease severity was assessed using the ALSFRS-R. Sleep quality, daytime sleepiness, and REM sleep behaviour disorder symptoms were evaluated using the PSQI, ESS and RBDQ questionnaires, respectively. Genetic analysis included targeted sequencing of *SOD1*, *TARDBP*, and *ANG*, followed by repeat expansion analysis of *C9orf72, ATXN1*, *ATXN2*, *ATXN3*, and *ATXN7*. Whole exome sequencing was performed in selected cases to further investigate the genetic basis of ALS.

**Results:** The cohort demonstrated a male predominance and an earlier age of onset compared to many Western populations. Poor sleep quality and excessive daytime sleepiness were frequently observed although no significant association was found between sleep parameters and disease severity. Genetic screening identified a pathogenic/likely pathogenic *TARDBP* variant classified by ClinVar while no known or novel pathogenic variants were detected in *SOD1* or *ANG*. Repeat expansion studies revealed one patient harbouring pathogenic *C9orf72* repeat expansions. For the *ATXN1*, one patient harboured pathogenic repeat expansions and two patients carried intermediate length repeats. Additionally, one patient harboured intermediate repeat expansions in *ATXN2*. Whole exome sequencing revealed novel and rare variants in ALS associated genes including *ANXA11, SIGMAR1, SETX, ERBB4 and NEK1,* highlighting substantial genetic heterogeneity.

**Conclusions:** Our findings highlight the complex genetic heterogeneity of ALS, with pathogenic repeat expansions and rare variants identified across multiple disease associated genes. These results demonstrate the importance of comprehensive genetic testing to improve the detection of clinically relevant variants in ALS. In addition, the high prevalence of sleep related disturbances observed in the cohort emphasizes the need to incorporate non motor assessment into routine clinical evaluation of patients with ALS.

## 1. Introduction

Amyotrophic Lateral Sclerosis (ALS), is a fatal neurodegenerative disease of the motor neuron network, marked by the progressive degeneration of upper and lower motor neurons. This neuronal loss leads to progressive muscle weakness, impaired mobility, and loss of essential functions causing complete paralysis over the years and untimely death due to respiratory failure(Goutman et al. 2022). ALS is considered a multifactorial disease, wherein both environmental factors and genetic predisposition, along with their complex interactions, play critical roles in disease onset and progression(Vasta et al. 2022). ALS is predominantly a late onset disorder, typically manifesting around the fifth or sixth decade of life, however considerable variability in the age of onset has been reported, including earlier presentation in certain geographical populations, especially Asians compared to their western counterparts(Lin et al. 2020). Several epidemiological studies have consistently indicated a male predominance, with a male to female ratio of approximately 2:1(Chio et al. 2020). Approximately 90-95% of the cases are sporadic, with no apparent family history, whereas the remaining 5-10% are familial and typically associated with inherited pathogenic variants in specific genes. However, advances in genetics have shown that the distinction between sporadic and familial ALS is not absolute, as genetic susceptibility also contributes to many apparently sporadic cases. Consequently, ALS is now recognised as a genetically heterogeneous disorder, with pathogenic variants in multiple genes disrupting diverse biological pathways that ultimately converge progressive motor neuron degeneration (Al-Chalabi, Andrews, and Farhan 2024; Menge, Decker, and Freischmidt 2025).

The widespread use of next generation sequencing has greatly expanded the number of genes linked to ALS. Variants in *C9orf72*, *SOD1*, *TARDBP*, and *FUS* account for substantial proportion of genetically resolved cases, while many additional genes contribute less frequently. Importantly, the distribution of these variants differs across populations. For example, *C9orf72* repeat expansion are the predominant genetic cause of ALS in European populations, whereas *SOD1* variants are encountered more often in Asian cohorts (Y.-P. Chen et al. 2021; Nishiyama et al. 2017; Z. Y. Zou et al. 2017). These differences highlight the need for population specific studies, particularly in underrepresented populations where the genetic spectrum of ALS remains incompletely defined.

Currently, there is no cure for the disease. The FDA approved pharmacological treatments-Riluzole, and Edaravone offer only modest benefits by extending the life expectancy by a few months (Jaiswal 2019). Recently, the FDA has approved Tofersen, the first antisense oligonucleotide drug targeting specifically for patients with *SOD1* mutations (Miller et al. 2020), marking a significant advancement in mutation specific treatment. One of the reasons that makes the diagnosis of ALS and thereby therapeutic intervention extremely difficult is the heterogeneous clinical and genetic presentation of the disease which varies substantially between individuals.

ALS typically presents with an insidious onset, often beginning with focal limb weakness in one of the limbs in spinal onset cases, which gradually progresses to involve other regions, including bulbar muscles, leading to dysarthria and dysphagia. In bulbar onset ALS, the disease initially manifests as subtle speech disturbances or swallowing difficulties, followed by progression to limb weakness and eventual loss of ambulation. Some individuals may also exhibit cognitive decline and behavioural impairments with signs showing emotional vulnerability, thereby falling in the ALS-FTD (Frontotemporal dementia) spectrum (Chio et al. 2020; Rusina, Vandenberghe, and Bruffaerts 2021; Strong et al. 2017; Ziser et al. 2025).

Clinical diagnosis of ALS is primarily based on neurological evaluation, supported by electromyography (EMG) and nerve conduction velocity (NCV) studies to rule out other ALS mimics(Vidovic et al. 2023). Diagnosis is further classified as definite, probable, or possible ALS according to the revised El Escorial criteria (Ludolph et al. 2015).

Since identifying *SOD1* through linkage analysis and candidate gene screening nearly three decades ago (Rosen et al. 1993), recent advances in genetic technologies, particularly next-generation sequencing, have led to the discovery of over 40 candidate genes associated with ALS (Suzuki et al. 2022). These findings have significantly expanded our understanding of the disease’s heterogeneous molecular aetiology, with ongoing research continuing to uncover additional genetic contributors.

While recent studies have improved our understanding of the genetic basis of ALS in Indian populations and have identified several diseases associated variants across different patient cohorts (Khurana et al. 2026; Narain et al. 2018; Vats et al. 2015, 2017). However, ALS is not defined by its genetics alone. Patients often show striking differences in disease onset, progression, symptom profile, and overall clinical course, even when carrying variants in the same gene (Tzeplaeff et al. 2024). This variability suggests that understanding ALS requires not only the identification of genetic factors but also careful clinical characterization of affected individuals.

In recent years, it has become increasingly clear that ALS extends beyond the selective degeneration of motor neurons. Alongside progressive muscle weakness and functional decline, many patients experience non motor symptoms that substantially affect daily life and quality of life. Among these, sleep related disturbances are particularly common. Poor sleep quality, frequent nocturnal awakenings, excessive daytime sleepiness, and respiratory difficulties during sleep have all been reported in ALS patients(Ahmed et al. 2015; Boentert 2019). Despite their clinical importance, these symptoms often receive less attention than motor manifestations, and their relationship with disease characteristics remains incompletely understood.

With this background, the present study sought to examine ALS from both a genetic and clinical perspective. Patients were screened for established ALS associated genes and repeat expansions, while WES was performed in selected genetically unresolved cases. In parallel, disease severity was assessed using the ALSFRS-R, and sleep related symptoms were evaluated using validated questionnaires. By combining genetic analysis with detailed clinical and sleep assessments within the same cohort, this study aimed to provide a broader picture of disease heterogeneity and to better understand the diverse factors that contribute to the ALS phenotypes.

## 2. Materials and methods

### 2.1. Recruitment of study participants

A total of 73 patients diagnosed with ALS were recruited from Department of Neurology, Institute of Medical Sciences, Banaras Hindu University, a tertiary care academic medical hospital in Northern India. Diagnosis was established by experienced neurologists according to the revised El Escorial criteria following comprehensive clinical evaluation and supporting investigations, including magnetic resonance imaging (MRI), electrophysiology (EMG), and nerve conduction velocity (NCV) studies. Detailed demographic and clinical information, including family history, was collected for each participant. Disease severity was evaluated using the Amyotrophic Lateral Sclerosis Functional Rating Scale-Revised (ALSFRS-R). Sleep related symptoms were assessed using the Pittsburgh Sleep Quality Index (PSQI), Epworth Sleepiness Scale (ESS), and REM Sleep Behaviour Disorder Screening Questionnaire (RBDSQ). Individuals presenting with symptoms suggestive of ALS but lacking confirmation through clinical and neurophysiological investigations were excluded from this study. Age and sex matched healthy controls were additionally screened for rare or novel variants identified during genetic analysis. The study was conducted following approval from the Institutional Ethics Committee, and written informed consent was obtained from all participants and/or their family members.

### 2.2. Genetic Studies

5 ml peripheral blood was collected in a EDTA vial from each patient after proper written informed consent and subsequently genomic DNA was extracted using salting out method. DNA was quantified using Nanodrop spectral analysis (Thermo Fischer, USA) and its quality was assessed by agarose gel electrophoresis (100V, 50 mins, 0.8% agarose gel in 1X Tris-acetate-EDTA buffer).

Targeted genetic screening of ALS candidate genes (*SOD1*, *TARDBP* and *ANG)*: Initially, mutation screening was done for 3 major ALS candidate genes including *SOD1, TARDBP,* and ANG, respectively. For that, 100ng of extracted genomic DNA was amplified using a thermal cycler (Applied Biosystems, USA) by PCR. The reaction mixture contained dNTPs, PCR buffer, 10 pmol/μL each of forward and reverse primers, Taq polymerase, and the volume was made up to 25μL using MilliQ water. The PCR product was purified by using Exonuclease1 (Exo1) and Shrimp Alkaline phosphatase (SAP) (USB Affymetrix, USA). Purified PCR product was subjected to cycle sequencing using BDTv.1.1 (Applied Biosystem, USA) and post-sequence cleanup, the product was run on a 3130 or 3500 ABI genetic analyzer (Applied Biosystems). Sequencing results were analyzed using Finch TV software (v.1.4.0) and the obtained sequences were compared with reference sequences using the NCBI BLAST tool.

Screening of repeat expansion of *C9orf72*: The samples were tested for the presence of pathogenic G4C2 hexanucleotide expansion in the first intron of C*9orf72* gene using repeat-primed PCR and fragment analysis using an ABI 3500 Dx genetic analyser (Applied Biosystems, USA) according to the protocol as previously described by Shamim et al. 2019. PCR conditions and the primers used in the study can be made available upon request.

Screening of *ATXN1, ATXN2, ATXN3 and ATXN7*: Trinucleotide repeat expansions in the *ATXN1*, *ATXN2*, *ATXN3*, and *ATXN7* were also screened using repeat-primed PCR and fragment analysis using an ABI 3500 Dx genetic analyser (Applied Biosystems, USA) according to the protocol previously established by Sharma et al. 2022).

### 2.3. Whole exome sequencing

Whole exome sequencing (WES) covering approximately 99% of coding regions and complete mitochondrial genome sequencing were performed using Illumina next generation sequencing platforms. Exome sequencing achieved an average coverage of 50-60×, while the mitochondrial genome was sequenced at a depth exceeding 1000X, with more than 90% of target bases covered at ≥ 20X. Sequence reads were aligned to the human reference genome (GRCh37/hg19) using BWA-MEM, and variant calling was performed following the GATK best practice workflow. Read duplication, base quality recalibration, and indel realignment were carried out using Sentineon tools, and variant were identified using the Sention HaplotypeCaller (Freed et al. 2017). A secondary variant calling pipeline based on DeepVariant was also employed for validation. Quality control filters were applied to exclude low confidence variants, and annotated variant datasets were generated using publicly available databases including OMIM, gnomAD, 1000 Genomes, GWAS Catalog, and MITOMAP.

Variants identified in the VCF file were prioritized based on their population frequency, sequencing quality and predicted functional impact. Variants with a minor allele frequency (MAF) below 1% or absent from population databases including 1000 Genome and gnomAD, were retained for further analysis. A minimum read depth of 20 and a CADD score of ≥20 was applied as additional filtering criteria. The potential functional effects of the prioritized variants were subsequently assessed *in silico* using SIFT, PolyPhen-2, PROVEAN prediction tools.

### 2.4. Statistical analysis

Data were analysed using appropriate descriptive and inferential statistical methods. Continuous variables were expressed as mean ± standard deviation (SD) while categorical variables were summarized as frequencies (%). Associations between categorical variables were assessed using Chi square test. Comparisons between two independent groups were performed using Mann Whitney U test or unpaired Student’s *t* test, depending on the distribution of data. Relationship between clinical and sleep related parameters were evaluated using correlation analysis where appropriate. All statistical tests were two tailed, and a *p* value < 0.05 was considered statically significant.

## 3. Results

### 3.1. Clinical features of the patients with novel variations

A total of 73 patients meeting the revised El Escorial Criteria for definite or clinically probable ALS were enrolled at the Institute of Medical Sciences, BHU, a tertiary referral centre in Northern India. The cohort predominantly comprised individuals from Northern India, with the majority originating from the Eastern regions of Uttar Pradesh, Bihar, and Chhattisgarh. The recruited patients were predominantly males (65.7%), with females comprising 34.2% reflecting a male: female ratio of 2:1 consistent with other population studies. The mean age of onset for all the patients was in the later years of the fourth decade (49.84 ± 10.37). Males exhibited a slightly earlier age of onset (48.54 ± 9.89) as compared to females (52.92 ± 11.13). Age wise distribution showed that, out of 73 patients, 23 patients (31.50%) were ≤ 44 years including 17 males and 6 females thereby indicating a young onset of ALS, 22 patients (30.13%) were between 45 and 54 with 16 males and 6 females, 22 patients (30.13%) were between 55 to 64 years with 12 males and 10 females and 6 patients (8.21%) were aged 65 years and above with 2 males and 4 females. Therefore, the age of onset was earlier than 50 years in 58.3% (28/48) in males and 44% (11/25) in females. There were no significant differences in the mean age between male and female patients. The earliest disease onset was observed in a male patient before 25 years of age, whereas the youngest female patient had onset before 30 years of age. About 72.60% of the patients diagnosed had spinal onset, while 27.39% patients had bulbar onset. The mean age of the bulbar onset patients was 52.85±10.39, while that of limb onset was 48.98 ±10.40 (**Table 1**) and there was no female predominance in the case of bulbar onset in our cohort (male: female 11:9) (**Table 2**).

**Table 1.**
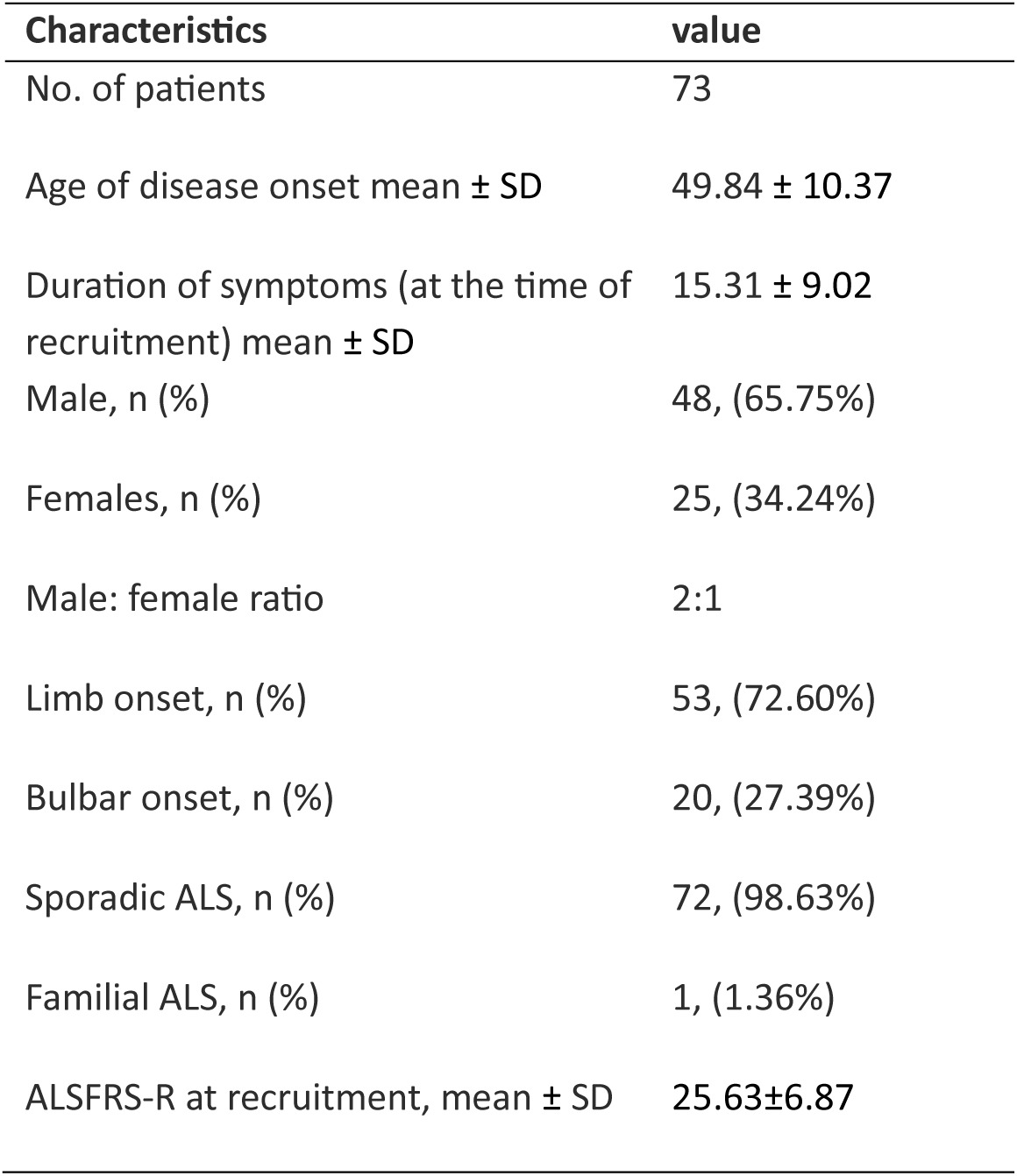
Demographic and clinical characteristics of the ALS cohort.

**Table 2.**
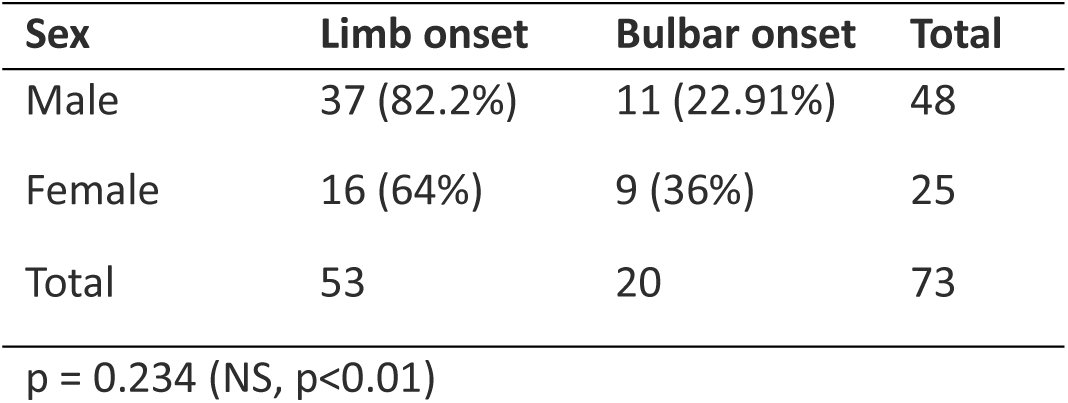
Association between sex and site of disease onset.

| **Sex** | **Limb onset** | **Bulbar onset** | **Total** |
|---|---|---|---|
| Male | 37 (82.2%) | 11 (22.91%) | 48 |
| Female | 16 (64%) | 9 (36%) | 25 |
| Total | 53 | 20 | 73 |

p = 0.234 (NS, p<0.01)

### 3.2. ALSFRS-R assessment

The mean ALSFRS-R recorded was 25.63±6.92. Based on functional status, six patients (8.21%) had mild functional impairment (ALSFRS-R: 37-48), thirty-four patients (46.57%) had moderate impairment (ALSFRS-S 25-36), and thirty-three patients (45.20%) showed severe functional impairment (ALSFRS-R ≤24). There were no statistically significant differences between ALSFRS-R between the two groups of patients (limb onset vs. bulbar onset).

### 3.3. Sleep characteristics of the ALS cohort

The mean ESS, PSQI, and RBDSQ scores were 9.78 ± 3.83, 10.91 ± 3.41, and 1.52 ± 1.60 respectively **(Table 3)**. Excessive daytime sleepiness (ESS ≥10) was observed in 40 patients (54.79%), while poor sleep quality (PSQI>5) was present in 67 (91.7%). Symptoms suggestive of REM sleep behaviour disorder (RBDSQ ≥5) were identified in 5 (6.84%) patients **(Table 4).**

**Table 3.** Sleep characteristics of ALS patients.

| Parameter | Mean | Range |
|---|---|---|
| ALSFRS-R | 25.63±6.92 | 12-42 |
| Epworth sleepiness scale (ESS) | 9.78 ± 3.83 | 2-17 |
| Pittsburgh Sleep quality Index (PSQI) | 10.91 ± 3.41 | 4-19 |
| RBD Screening Questionnaire (RBDSQ) | 1.52 ± 1.60 | 0-7 |

**Table 4.** Prevalence of sleep disturbances in ALS patients.

| **Parameter** | **Cut-off** | **n (%)** |
|---|---|---|
| Excessive daytime sleepiness | ESS ≥ 10 | 40 (54.79%) |
| Poor quality sleep | PSQI >5 | 67 (91.7%) |
| Probable REM sleep behaviour disorder | RBDSQ ≥ 5 | 5 (6.84%) |

No significant correlation was observed between ALSFRS-R score and ESS, PSQI or RBDSQ scores (p>0.05) (**Table 5**). Similarly, comparison of limb onset and bulbar onset patients revealed no significant differences in ALSFRS-R, ESS, PSQI, or RBDSQ score (p>0.05 for all comparisons) **(Table 6).**

**Table 5.**
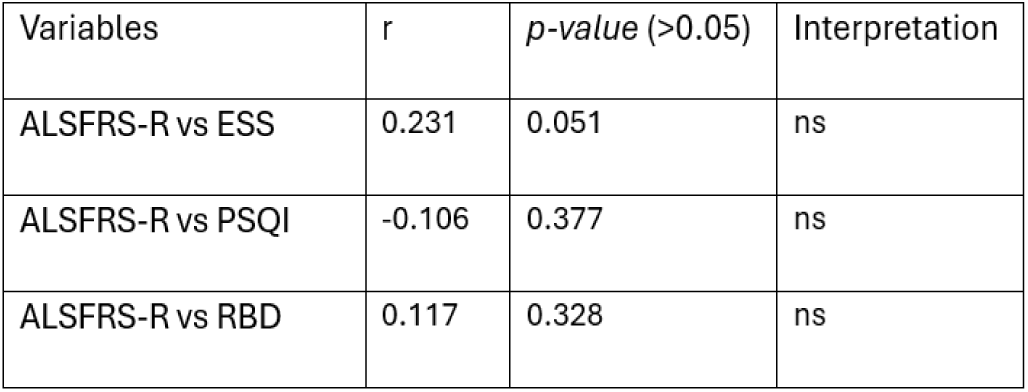
Relationship between disease severity and sleep-related measures.

| Variables | r | *p-value* (>0.05) | Interpretation |
|---|---|---|---|
| ALSFRS-R vs ESS | 0.231 | 0.051 | ns |
| ALSFRS-R vs PSQI | -0.106 | 0.377 | ns |
| ALSFRS-R vs RBD | 0.117 | 0.328 | ns |

**Table 6.** Comparison of clinical and sleep parameters between limb onset and bulbar onset ALS patients.

| **Variable** | **Limb onset n=53** | **Bulbar onset n=20** | **p-value (Two tailed)** |
|---|---|---|---|
| ALSFRS-R | 26 ±7.24 | 24.7± 6.05 | 0.47 |
| ESS | 10.07 ± 4.01 | 9 ± 3.27 | 0.24 |
| PSQI | 11.09 ± 3.33 | 10.45 ± 3.67 | 0.50 |
| RBDSQ | 1.49 ± 1.67 | 1.6 ± 1.46 | 0.78 |

### 3.4. Targeted genetic screening of candidate genes

Targeted gene screening by Sanger sequencing of all five exons of *SOD1* revealed a potential polymorphism; **p. Ser35Ile** (**Fig 1, Table7**) however no known or novel pathogenic variants were identified by screening *SOD1* or the coding exon of *ANG* in our cohort.

**Fig 1.**
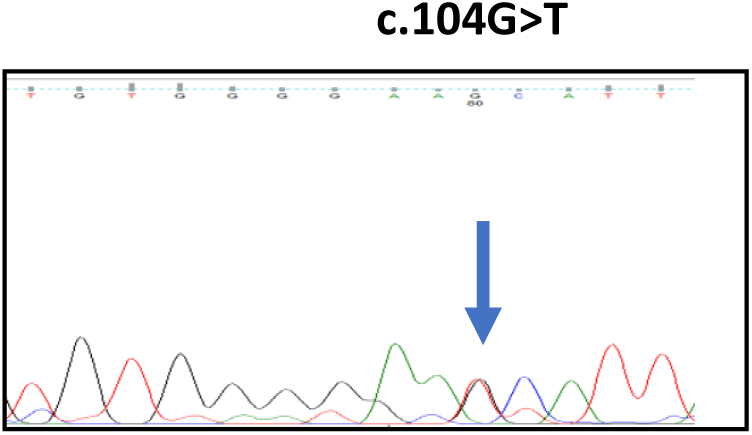
Sanger sequencing chromatogram showing the identification of the variant p. Ser35Ile in *SOD1.* Representative chromatogram of variant sequences with respective nucleotide substitutions indicated by arrows.

Analysis of *TARDBP* identified a heterozygous variant p.Ile383Val (NM_007375.4; c.1147A>G; rs80356740) in one patient (PS 18) (**Fig 2**, **Table 7**). This variant is classified as likely pathogenic in ClinVar and previously reported in Greek FTD patients (Charoniti et al. 2021). The carrier was a male patient in the 45-54 years of age group presenting with spinal onset ALS without bulbar involvement and a baseline ALSFRS-R score of 26. No additional pathogenic variants were identified in this gene.

**Fig 2.**
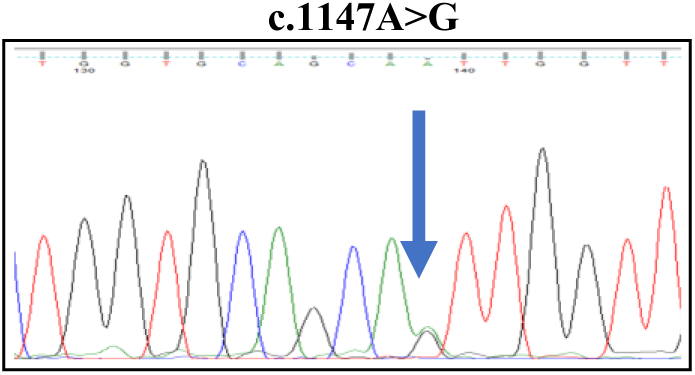
Sanger sequencing chromatogram showing the identification of the variant p.Ile383Val in *TARBDP.* Representative chromatogram of variant sequences with respective nucleotide substitutions indicated by arrows.

**Table 7.**
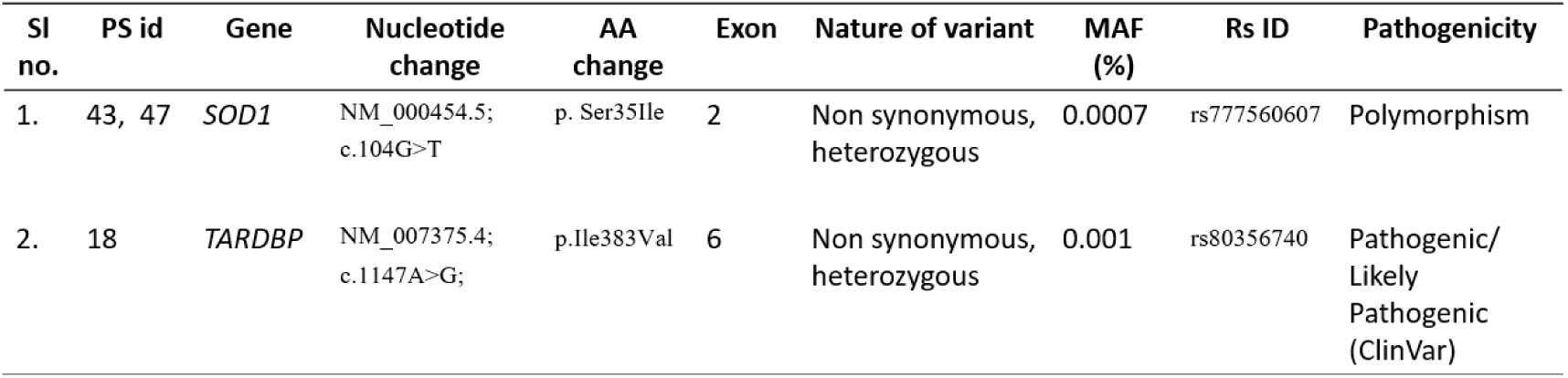
Summary of variants identified in the ALS cohort by Targeted Sequencing of selected ALS genes.

| Sl no. | PS id | Gene | Nucleotide change | AA change | Exon | Nature of variant | MAF (%) | Rs ID | Pathogenicity |
|---|---|---|---|---|---|---|---|---|---|
| 1. | 43, 47 | *SOD1* | NM_000454.5; c.104G>T | p. Ser35Ile | 2 | Non synonymous, heterozygous | 0.0007 | rs777560607 | Polymorphism |
| 2. | 18 | *TARDBP* | NM_007375.4; c.1147A>G; | p.Ile383Val | 6 | Non synonymous, heterozygous | 0.001 | rs80356740 | Pathogenic/Likely Pathogenic (ClinVar) |

### 3.5. Repeat Expansion Analysis

Repeat primed PCR screening of *C9orf72*, *ATXN1*, *ATXN2*, *ATXN3*, and *ATXN7* identified one female patient (PS 05) harbouring a pathogenic *C9orf72* expansion exceeding the established disease threshold (>32 repeats). She presented with limb onset ALS that progressed to bulbar involvement over time, had a disease duration of 24 months, no significant family history, and demonstrated an ALSFRS-R score of 25 at evaluation. The patient also exhibited prominent sleep related complaints, including disturbed nocturnal sleep and excessive daytime fatigue.

Compared with controls, patients exhibited a higher frequency of intermediate and pathogenic expansions in *ATXN1* and *ATXN2*. One individual (PS 43), a male patient within 55-64 years age group with a disease duration of 12 months and limb onset symptoms, carried a pathogenic *ATXN1* expansion of 42 repeats (disease threshold is >39 repeats). Two additional patients both within the age group of 45-54 years; PS 15 with limb onset symptoms and PS 51 with limb onset that subsequently progressed to bulbar involvement, exhibited intermediate *ATXN1* expansions (36 repeats each). An intermediate length *ATXN2* expansion (30 repeats, disease threshold ≥32 repeats) was identified in one female patient (PS 52) within the age group of 55-64 years with bulbar onset symptoms. No intermediate or pathogenic repeat expansions were detected in *ATXN3* or *ATXN7*.

### 3.6. Whole exome sequencing

A subset of patients was selected for whole exome sequencing (WES) based on disease severity, as assessed using the ALSFRS-R scores. The clinical characteristics of the patients included in the WES analysis are presented in **Table 8**. WES identified rare and potentially pathogenic variants in multiple established ALS candidate genes, including *SETX, ANXA11, SIGMAR1*, *ERBB4*, and *NEK1* (**Table 9**). In PS 57, a novel heterozygous, non synonymous, missense variant p.Val416Ala (c.1247T>C) in *ERBB4* was identified; this variant was absent from population databases and predicted to be deleterious by various in silico pathogenicity prediction tools. PS 6 harboured a rare heterozygous missense variant p. Gln1349Lys (c.4045C>A) in *SETX* (MAF 0. 000008), annotated as a variant of uncertain significance in ClinVar but consistently predicted to be damaging by various in silico pathogenicity prediction tools, along with a homozygous, non-synonymous variant c.*31A>G in the 3’UTR of *SIGMAR1*, predicted to be pathogenic. Notably, a novel *NEK1* variant p. Gln592X (c.1774C>T) classified as likely pathogenic according to ACMG guidelines was identified in PS 39. A young onset ALS patient was found to carry a rare variant p. Arg452Trp (c.1354C>T) in *ANXA11* together with a pathogenic variant in *SIGMAR1* p.Arg208Trp (c.622C>T, rs11559048) as reported previuosly (Roychowdhury et al. 2024).

**Table 8.** Clinical data for patients carrying novel and/or rare variants in the ALS candidate genes.

| Patient ID | Criteria | Gender | Age range (years) | Duration of symptoms | Place of Onset | Spread | ALSFR-R<br>(Max Score=48) |
|---|---|---|---|---|---|---|---|
| PS_04 | Definite ALS | Male | 35-44 | 12 months | Limb | -- | 19 |
| PS_06 | ALS+FTD | Female | 55-64 | 4 months | Limb | Bulbar | 22 |
| PS_28 | Definite ALS | Male | <25 | 14 months | Limb | Bulbar | 21 |
| PS_39 | Definite ALS | Female | <44 | 24 months | Limb | -- | 15 |
| PS_45 | Definite ALS | Male | 55-64 | 24 months | Bulbar | -- | 19 |
| PS_57 | Definite ALS | Male | >65 | 12 months | Limb | Bulbar | 18 |

**Table 9.** List of variants in ALS associated genes identified through WES in our cohort.

| Patient ID | Gene symbol | Chromosome | Zygosity | Nucleotide change | Amino Acid Change | Nature of change | MAF % | RS ID | ClinVar | In silico Prediction (SIFT, PolyPhen, Mutation taster) |
|---|---|---|---|---|---|---|---|---|---|---|
| **PS_57** | *ERBB4* | 2 | Heterozygous | c.1247T>C | p. Val416Ala | Non Synonymous | Novel | NR | NR | Pathogenic |
| **PS_06** | *SETX* | 9 | Heterozygous | c.4045C>A | p.Gln1349Lys | Non synonymous | 0.000008 (global) | NR | Uncertain Significance | Pathogenic |
|  | *SIGMAR1* | 16 | Homozygous | Non synonymous (A>G) | ----<br>(c.*31A>G) | 3'UTR | 0.92 (global) | rs4879809 | R | Pathogenic |
| **PS_39** | *NEK1* | 4 | Heterozygous | c.1774C>T | p.Gln592X | Nonsense | Novel | NR | NR | Pathogenic |
| **PS_28** | *ANXA11* | 10 | Heterozygous | (c.1354C>T) | p.Arg452Trp | Non synonymous | 0.00915 (Asian ) | -- | NR | Pathogenic |
|  | *SIGMAR1* | 16 | Homozygous | c.622C>T | p.Arg208Trp | Non synonymous | 0.007 (Global ) | rs11559048 | R | Pathogenic |

## 4. Discussion

ALS is among the most devastating and fatal neurodegenerative disorders, phenotypically characterized by progressive paralysis and untimely death, affecting individuals of all ethnicities worldwide. The disease is increasingly recognized as a clinically and biologically heterogeneous disorder, with considerable variability in both its genetic architecture and disease presentation (Tzeplaeff et al. 2024). While substantial progress has been made in identifying genes associated with ALS, the factors contributing to differences in clinical manifestations, particularly non motor symptoms, remain incompletely understood. Therefore, in the present study, we aimed to explore the genetic basis of 73 ALS patients from one of the largest tertiary care hospitals in North India by combining targeted genetic screening, repeat expansion analysis, whole exome sequencing (WES) technique, and clinical assessment of sleep disturbances.

The demographic characteristics of our cohort were largely consistent with previous epidemiological studies. Similar to reports from other populations, the incidence was higher in males than females, yielding a male-to-female ratio of 2:1, (Longinetti and Fang 2019) (Chio et al. 2020). This male predominance has been consistently observed across multiple ALS cohorts and may reflect complex interactions between genetic, hormonal, and environmental factors (Czlonkowska et al. 2005; Trojsi et al. 2020). The majority of the patients in the present study had sporadic ALS, with only a single patient reporting a possible positive family history. This observation is in agreement with the widely accepted view that sporadic forms constitute the largest proportion of ALS cases worldwide (Feldman et al. 2022; Wolfson et al. 2023).

An interesting finding was the relatively young age of disease onset (49.84± 10.37 years) observed within our cohort which is significantly lower (p<0.05) than the reported pool means of 62.1– 66.3 years old in European population (Al-Chalabi et al. 2014), (Sánchez-Martínez et al., 2019). A similar trend of young onset ALS cases has been seen in other Asian countries like Chinese population, where the mean age recorded was at 48-53 years (Gao et al., 2021), (Longinetti & Fang, 2019,),(Chen et al., 2021). Our findings align more closely with those reported from other Indian and East Asian populations, where disease onset frequently occurs in the later part of the fourth decade of life (Khurana et al. 2026; Nalini et al. 2008). Population based studies have found that individuals with fALS tend to develop the disease earlier than that of the sALS patients, and for the sALS patients, mostly they are affected in the fifth or sixth decade of their life (Marin et al. 2015), however sALS patients in our cohort have the disease onset much earlier in the fourth decade, than those reported in the American or Caucasian population, with the mean age of onset at 49.84 ± 10.37 years. Previous studies have found that females have a predominance of bulbar onset compared to males (He et al., 2022), (Chio et al. 2020) but our study did not reveal any such correlation, where the male: female ratio was at 11:9.

Beyond demographic characterization, the present study also examined disease severity and sleep related symptoms. The mean ALSFRS-R score suggested that most patients were recruited during moderate stages of disease progression, although considerable variation was observed across the cohort. Such heterogeneity is expected in a disease like ALS and reflects differences in disease duration, progression rate, site of onset, and individual disease trajectories.

Sleep disturbances emerged as a prominent non motor feature within the study population. Poor sleep quality was the most frequently reported abnormality, with more than half of the patients (91.7%) demonstrating PSQI scores indicative of clinically significant sleep impairment. Excessive daytime sleepiness was also observed in a substantial proportion of individuals (54.79%), whereas symptoms suggestive of REM sleep behaviour disorder were observed in 6.84% patients which was comparatively less common in our cohort. These findings support growing evidence that sleep dysfunction represents an important but often under recognized component of ALS. While clinical attention is traditionally focused on motor disability and respiratory decline, sleep disturbances can substantially affect quality of life, cognitive performance, emotional wellbeing, and caregiver burden as can be seen in other neurodegenerative disorders as well apart from ALS (Bartolomei et al. 2018; Boentert 2019).

The mechanisms responsible for impaired sleep in ALS are likely multifactorial. Progressive muscle weakness can make repositioning during sleep difficult, leading to repeated awakenings and fragmented sleep. Muscle cramps, pain, fasciculations, and nocturnal discomfort may further contribute to [poor sleep quality (Boentert 2019; Lucia et al. 2021). Respiratory muscle involvement represents another important factor, as nocturnal hypoventilation can develop before overt daytime respiratory symptoms become clinically relevant (Gnoni et al. 2023; Lucia et al. 2021). In addition, psychological stress, anxiety, and concerns regarding disease progression may further exacerbate sleep disturbances. The coexistence of multiple contributing factors likely explains the considerable variability in sleep related symptoms observed among ALS patients.

Interestingly, sleep disturbances appeared to occur across the disease spectrum rather than being confined to specific clinical subgroups. No significant differences were observed between limb onset and bulbar onset patients, nor where there were any significant differences between male and female patients (**Table 2**). Furthermore, ALSFRS-R scores did not have any significant correlations with ESS, PSQI, or RBDSQ scores (**Table 5**). Although weak trends were observed, sleep related symptom did not appear to be directly linked to functional status at the time of assessment. This suggests that conventional motor disability scales may not fully capture the factors responsible for sleep dysfunction in ALS. As mentioned earlier, respiratory involvement, mood disorders, medication effects, pain and individual susceptibility may all influence sleep independently of overall motor impairment the absence of significant associations should also be interpreted in light of the cross-sectional design of the study, as sleep disturbances and motor decline may not progress in parallel throughout the disease course.

The genetic findings of this study further highlight the complexity and heterogeneity of ALS. Comprehensive screening of *SOD1* and *ANG* did not identify pathogenic variants within the cohort. Although *SOD1* remains one of the most extensively studied ALS genes worldwide, its prevalence varies considerably among different populations. Similarly, *ANG* variants have been reported in a small proportion of ALS patients and are often considered uncommon contributors to disease susceptibility. The absence of pathogenic variants in these genes within our cohort may therefore reflect population specific genetic distributions.

Only a single patient carried a likely pathogenic *TARDBP* variant (rs80356740). *TARDBP* encodes TDP-43, an RNA binding protein involved in multiple aspects of RNA metabolism, including splicing, transport, and transcriptional regulation (Balendra et al. 2025). Pathological aggregation and cytoplasmic mislocalization of TDP-43 represent one of the defining pathological hallmarks of ALS (Bilican et al. 2012; Smethurst et al. 2020). Interestingly, the identified variant had been previously reported in a patient with frontotemporal dementia, further emphasizing the clinical and pathological overlap between the neurodegenerative disorders (Charoniti et al. 2021). Although only one *TARDBP* positive patient was identified, this finding reinforces the central role of RNA dysregulation in ALS pathogenesis.

In our cohort, a single patient was also found to carry a pathogenic *C9orf72* repeat expansion. While *C9orf72* expansions represent the most commonly known genetic cause of ALS in many European populations, they appear to be less frequent in Asian cohorts (Liu et al. 2018; Z.-Y. Zou et al. 2017) (Vats et al. 2017). Our finding is consistent with previous studies in Indian ALS patients, where *C9orf72* expansions have been reported in approximately 3.2% of patients and represent an established genetic contributor to disease susceptibility (Shamim et al. 2019; Vats et al. 2017a). Although the frequency of *C9orf72* expansions in India appears lower than that observed in several European cohorts, their identification remains clinically relevant. The detection of a repeat expansion in the present study further supports the inclusion of *C9orf72* testing in routine genetic screening of ALS patients particularly as such mutations cannot be identified through conventional exon based sequencing approaches.

In addition to *C9orf72* screening, repeat expansion analysis was performed in selected spinocerebellar-ataxia (SCA) associated genes; *ATXN1, ATXN2, ATXN3, ATXN7*. One clinically confirmed ALS patient was found to harbour an expanded pathogenic *ATXN1* allele (42 repeats), while two other patients carried intermediate lengths repeat expansions (36 repeats respectively) in the same gene. An intermediate length repeat expansion (30 repeats) in *ATXN2* was also identified in one patient in our cohort whereas no pathogenic expansions were detected in *ATXN3* or *ATXN7*.

The coexistence of ALS and a pathogenic range *ATXN1* expansion in the same individual appear to be rare. To the best of our knowledge, this is the first report of such an observation in an ALS patient. Although pathogenic *ATXN1* expansions are primarily associated with spinocerebellar ataxia type 1 (SCA1), intermediate length *ATXN1* repeats have been suggested to increase the susceptibility to ALS. Thus, the finding in our cohort is particularly noteworthy, although it remains unclear whether the pathogenic expansion contributed to the ALS phenotype or represents a coincidental finding. Similarly, intermediate *ATXN2* expansions are recognized as genetic risk factors for ALS and are thought to increase disease susceptibility rather than cause the classical SCA phenotype. The presence of intermediate length expansions in both *ATXN1* and *ATXN2* is in accordance with previous observations suggesting that repeat length variations in these genes may influence ALS susceptibility or disease expression (Novy et al. 2024). These findings also point to the possibility of overlapping clinical features among repeat associated neurodegenerative disorders highlighting the complex genetic architecture underlying motor neuron disease (Morello et al. 2020; Novy et al. 2024; Tazelaar et al. 2020). The absence of expansions in *ATXN3* and *ATXN7* further suggests that suggest that such repeat expansions are unlikely to represent common contributors to ALS susceptibility within the present cohort.

Whole exome sequencing was performed in eight patients with an ALSFRS-R score <25, who remained genetically unresolved following targeted screening. Although the number of cases analysed was limited, the findings provide additional insight into the genetic complexity of ALS. Rare and potentially pathogenic variants were identified in several genes previously implicated in ALS; including *ANXA11, SIGMAR1*, *SETX*, *ERBB4*, and *NEK1*. Notably, a number of these variants were absent or extremely rare in publicly available population databases, highlighting the possibility that rare variants may contribute to disease susceptibility in at least a subset of patients.

These genes participate in diverse biological pathways relevant to motor neuron survival. *ANXA11* has been implicated in vesicle trafficking and stress granule dynamics, processes increasingly recognized an important mechanism in neurodegeneration. *SIGMAR1* plays a critical role in endoplasmic reticulum signalling, calcium homeostasis, and cellular stress responses. *SETX* encodes senataxin, a helicase involved in RNA processing and DNA repair, while *ERBB4* contributes to neuronal development and synaptic signalling(Takahashi et al. 2013). *NEK1* has recently emerged as an important ALS susceptibility gene involved in DNA damage repair, microtubule organization, microtubule formation, ciliogenesis all processes that are essential for neuronal integrity(Martins et al. 2021; Shalom et al. 2008). Loss of function variants in *NEK1* have been repeatedly reported to increase the risk of ALS (Mann et al. 2023), making the identification of a novel truncating variant in our cohort noteworthy.

The identification of variants affecting multiple biological pathways supports the view that ALS is not a single gene disorder but rather a genetically heterogeneous condition arising from disruptions in interconnected cellular processes (Maurel et al. 2018). Importantly, several variants identified in the present study were absent from the publicly available population databases, suggesting that they may represent rare or population specific genetic alterations. Such findings underscore the importance of conducting genetic studies in underrepresented populations, as variants that are rare or absent in European cohorts may contribute significantly to disease risk elsewhere.

Another notable observation was the detection of multiple potentially relevant variants within individual patients as seen in a previously reported young onset ALS case harbouring both *ANXA11* and *SIGMAR1* variants (Roychowdhury et al. 2024). This finding supports the emerging concept that ALS may, in some cases have an oligogenic basis, whereby the cumulative effect of multiple genetic variants contributes to disease susceptibility and progression. Similar observations have been reported in other ALS cohorts (Lattante et al. 2019; McCann et al. 2021).

Taken together, the findings of this study highlight heterogeneity in terms of both clinical presentation and genetic profiles of ALS in the Indian population. Sleep disturbances were common and represented an important non motor aspect of the disease, yet they were not directly associated with functional disability or clinical subtype. Simultaneously, genetic analyses revealed a diverse spectrum of novel, rare and potentially pathogenic variants, many of which affected distinct but biologically convergent pathways involved in neuronal survival. While the contribution of several variants requires further validation, their identification highlights the considerable genetic heterogeneity underlying ALS. As larger sequencing datasets and functional studies become available, improved interpretation of rare variants will enhance our understanding of disease mechanism. Such efforts will be important not only for improving variant interpretation but also for refining molecular classification of ALS and identifying patient subgroups that may benefit from future targeted therapeutic approaches. Collectively, these results emphasize the need for integrated and clinical and genetic investigations to better capture the complexity of ALS.

## Acknowledgement

The authors gratefully acknowledge the patients and their family.

## Statement of Ethics

This study protocol was reviewed and approved by Institutional Ethics Committee, Institute of Science, Banaras Hindu University,Varanasi, India (Ref No: I.Sc/ECM-XII/2021-22).

## Conflicts of Interests

The authors have no conflicts of interests

## Funding Sources

First Author acknowledges DST Inspire for providing fellowship.

## Author Contributions

**SRC:** Conceptualization, Methodology, Formal analysis, Validation, Investigation, Writing-original draft. **PD:** Conceptualization, Writing-review & editing, Resources, Supervision, Project administration, Funding acquisition.**DJ:** Methodology, Resources, Supervision, Project administration, **AK:** Resources

## Data Availability Statement

The datasets generated during and/or analysed during the current study are available from the corresponding author upon reasonable request.

## Notes

### Competing Interest Statement

The authors have declared no competing interest.

